# Protocol for piloting a phone-based web survey to elicit barriers to accessing triage camps within community-based, Peek-powered eye screening programmes

**DOI:** 10.1101/2023.06.15.23290866

**Authors:** Luke Allen, Sailesh Mishra, Malebogo Tlhajoane, Bakgaki Ratshaa, Dave Macleod, Hannah Chroston, Oathokwa Nkomazana, Andrew Bastawrous

## Abstract

**Title:** Piloting a phone-based web survey to elicit barriers to accessing triage camps within community-based, Peek-powered eye screening programmes

**Design:** Pilot test of an online survey sent via SMS to 1,600 parents/guardians of children who did not attend eye clinics having been found to have an eye need at school screening. We will perform the same pilot twice; once in Botswana and once in Nepal

**Aims:** To quantify the response rate among 16 different subgroups of recipients according to gender, location, wealth and assets.

Note that we are not concerned with the responses at this pilot phase – only the suitability of the medium.

**Population:** Parents/guardians of children who did not attend screening within Botswana’s Peek-powered national school-based ‘Pono Yame’ programme, and participants in Nepal’s Peek-powered community screening programme

**Intervention:** A four-question online survey exploring barriers to attendance. The hyperlink will be sent via SMS. Non-responders will receive a single follow-up reminder 7 days later.

**Outcome measure:** Cumulative survey completion rates over the 31 days that follow the original SMS invitation.

**Duration:** All survey invitations will be sent on the same day. As such, the pilot will last 31 days.

## Background

### Global eye service attendance

Peek eye screening programmes operate all over the world. Hundreds of thousands of people have their vision checked every year, and approximately a third of everyone who is seen requires some form of treatment. Unfortunately, only around half of those identified as needing further care show up at the clinic or hospital. Women, widows, and those living in rural areas appear to be the least likely to attend, however we don’t fully understand the reasons why, or how to increase attendance rates among these subgroups.

This is a ubiquitous problem: beyond eye screening, attendance rates for all types of medical clinic are <50% across Africa. One-off qualitative projects are occasionally undertaken to explore potential barriers, but these are time-consuming, expensive, and often produce findings with limited generalisability. Online surveys are a relatively cheap and rapid way of obtaining data, however it is likely that the most marginalised groups in low and middle income countries may not have access to a phone or sufficient online data.

### Nepal attendance

This Peek-based screening programme based in and around Lahan is in its infancy, with approximately 300 people screened so far. To date, internal data suggest that less than a third of those referred attend for triage and treatment.

### Pono Yame attendance

To date, 55,000 schoolchildren have been screened in the Pono Yame programme. Five thousand of these children (9%) have been referred to triage and treatment camps, however only 1/3 have attended.

### Rationale

If the numbers we have seen so far hold across both programmes, then many people will not receive the care they need. It is expected that the entire Pono Yame programme will screen 500,000 children, of whom 45,000 will be identified with an eye need at screening and referred to triage and treatment camps. If the attendance rate is not boosted from 33%, then 30,000 children will not receive the care they need.

It is imperative that we understand the barriers to access for these children, so that we can modify the programme to boost attendance.

Our continuous improvement team are planning to work with (parents of/) non-attenders to understand the main barriers to access and identify potential solutions. In 2023 our team will conduct interviews with approximately 30 parents, teachers, and screeners to identify the main barriers to access in both countries. To check whether the resulting list of barriers is representative, we want to send out a short survey to a much larger sample non-attenders so that they can rank the barriers in order of perceived importance. We want to maximise validity, speed and response rates whilst minimising costs as we are trying to encourage global health funders to take on this work once/if we can show that engaging with marginalised groups leads to higher access rates.

There is evidence that short online surveys (including those sent or administered via SMS) provide high quality, reliable data at low cost and with good speed. Clearly there is an issue with mobile phone coverage and data access in rural and less affluent parts of Nepal and Botswana that make this approach challenging. However, social enterprises like ‘Play Verto’ have used similar data-led community engagement to achieve response rates of >80% across a range of low- and middle-income countries. We also note that Nepal and Botswana have high smartphone penetration, with approximately 100 mobile phone subscriptions for every 100 people.

### Aim

In this pilot study we aim to quantify the response rate for a short online survey, sent to parents of non-attenders via a hyperlink in an SMS. We already have the mobile numbers for every non-attender, as these are gathered during screening.

At this stage we are interested in the response rate among different groups – particularly among rural and low-income groups that are likely to have the lowest access to smart phones and reliable coverage.

## Process of developing the pilot survey

Previous Peek work has identified seven common types of barriers to accessing eye services:

- **A**wareness
- **B**ad services - e.g., unstaffed, unstocked
- **C**osts
- **D**istances
- **E**scorts - does the service accommodate them?
- **F**ear - e.g., myths around eye surgery
- **G**ender

The Peek CEO, our Nepalese and Batswana Research Coordinators, and the LSHTM-based Research Manager have developed a list of ten questions aligned with these domains:

- **A**wareness
  ▪ It’s not that important to me
- **B**ad services - e.g., unstaffed, unstocked
  ▪ The staff are rude or poorly trained
  ▪ The clinic does not have enough supplies
  ▪ The clinic is not open at the time I want to go
- **C**osts
  ▪ I can’t afford to go
  ▪ I can’t afford the treatment
- **D**istances
  ▪ The clinic is hard to travel to
- **E**scorts - does the service accommodate them?
  ▪ My child or I need an escort/chaperone to attend
- **F**ear - e.g., myths around eye surgery
  ▪ I’m worried about the treatment
- **G**ender
  ▪ My child’s gender makes things difficult

We translated these into Nepalese and Setswana. Appendix 1 provides the full instrument in English. The Play Verto team turned the questions and response options into an attractive online survey ([here](https://client.playverto.com/peek-vision/botswana-adult)).

In each country we will send the hyperlink via SMS to a sample of 1,600 parents/guardians: 100 from each of the following 16 sociodemographic groups:

Male - Concrete floor (proxy for higher wealth)

- male, concrete floor, urban, own a phone
- male, concrete floor, urban, associated phone number does not belong to parent/guardian
- male, concrete floor, rural, owns a phone
- male, concrete floor, rural, associated phone number does not belong to parent/guardian

Male - Other floor type (proxy for lower wealth)

- male, other floor type, urban, own a phone
- male, other floor type, urban, associated phone number does not belong to parent/guardian
- male, other floor type, rural, owns a phone
- male, other floor type, rural, associated phone number does not belong to parent/guardian

Female - Concrete floor (proxy for higher wealth)

- female, concrete floor, urban, own a phone
- female, concrete floor, urban, associated phone number does not belong to parent/guardian
- female, concrete floor, rural, owns a phone
- female, concrete floor, rural, associated phone number does not belong to parent/guardian

Female - Other floor type (proxy for lower wealth)

- female, other floor type, urban, own a phone
- female, other floor type, urban, associated phone number does not belong to parent/guardian
- female, other floor type, rural, owns a phone
- female, other floor type, rural, associated phone number does not belong to parent/guardian

The hyperlink will be sent via SMS. Non-responders will receive a single follow-up reminder 7 days later.

## Outcomes

If the response rate is >50% for each group then we will take the approach to scale. As such, our primary outcome is response rate (all questions completed) for people with each characteristic at day 7. We will take the approach to scale if the bottom of the 95% confidence interval is >50% for each group.

The secondary outcomes are:

- Response rate at day 31.
- Overall response rate at days 7 and 31.
- Mean time to response within each group
- Proportion of people who complete at least one of the items but not all of them.

## Analysis

We report the proportion of people that complete the survey with each of the characteristics.

## Consent

This survey contains four questions. The instrument carries negligible risks to physical or mental health. As such, the requirements for informed consent differ from what is required for an interventional trial; more intrusive or onerous forms of data collection; or activities that elicit sensitive information.

In line with other negligible-risk SMS surveys, we will obtain implied consent i.e. the process of opening the link and completing the survey will be taken as evidence of consent.^1^

The participant information (Appendix 1) will be presented to users on the landing page that says ‘‘Essential participant information: who we are and how we use your data”. Clicking on this text displays the full PIL. The technology provider (Play Verto) does not currently enable tick boxes, so we will take survey completion as consent.

Our invitation SMS is 160 characters (i.e. 1 SMS) long to minimise the burden on potential respondents. The wording is brief and direct, but manages to convey why we are seeking data, and the fact that this is a polite, optional request rather than a mandatory activity. Participants will have received previous messages from the same number providing appointment details. The landing page and survey uses images from screening activities to remind potential responders who we are. The follow-up information on the landing page further reinforces that we will use the data to make improvements to the programme:

### SMS message

“We noticed your child didn’t attend for eye care after screening

Please complete this short survey to tell us why

*https://tinyurl.com/4xd8bvz5*

Thanks!”

### Survey landing page wording

*“Hi! We noticed that you did not make it to the clinic. We want to understand if there is anything we could do differently that would help you and others like you get the care you need*.*”*

It is very important that we minimise the amount of text sent my SMS and on the landing page so that we keep the cognitive burden to an absolute minimum.

## Output and next steps

We will use the findings internally to assess whether online surveys are a feasible tool to use for our next research project: <u>eliciting barriers and solutions to improve clinic access</u>. If the response rate is >50% for all then we will consider taking this SMS-based online survey to scale, using it alongside in-person and/or telephone interviews to gather data on the barriers faced by different groups.

If we find that the response rate is particularly low for one or two groups, or intersectional subgroups (e.g. rural women who don’t own a phone) then we could consider using SMS online surveys for all other groups but alternate methods for this subgroup.

## Data Availability

All data produced in the present work are contained in the manuscript.

## Appendix 1: Participant information

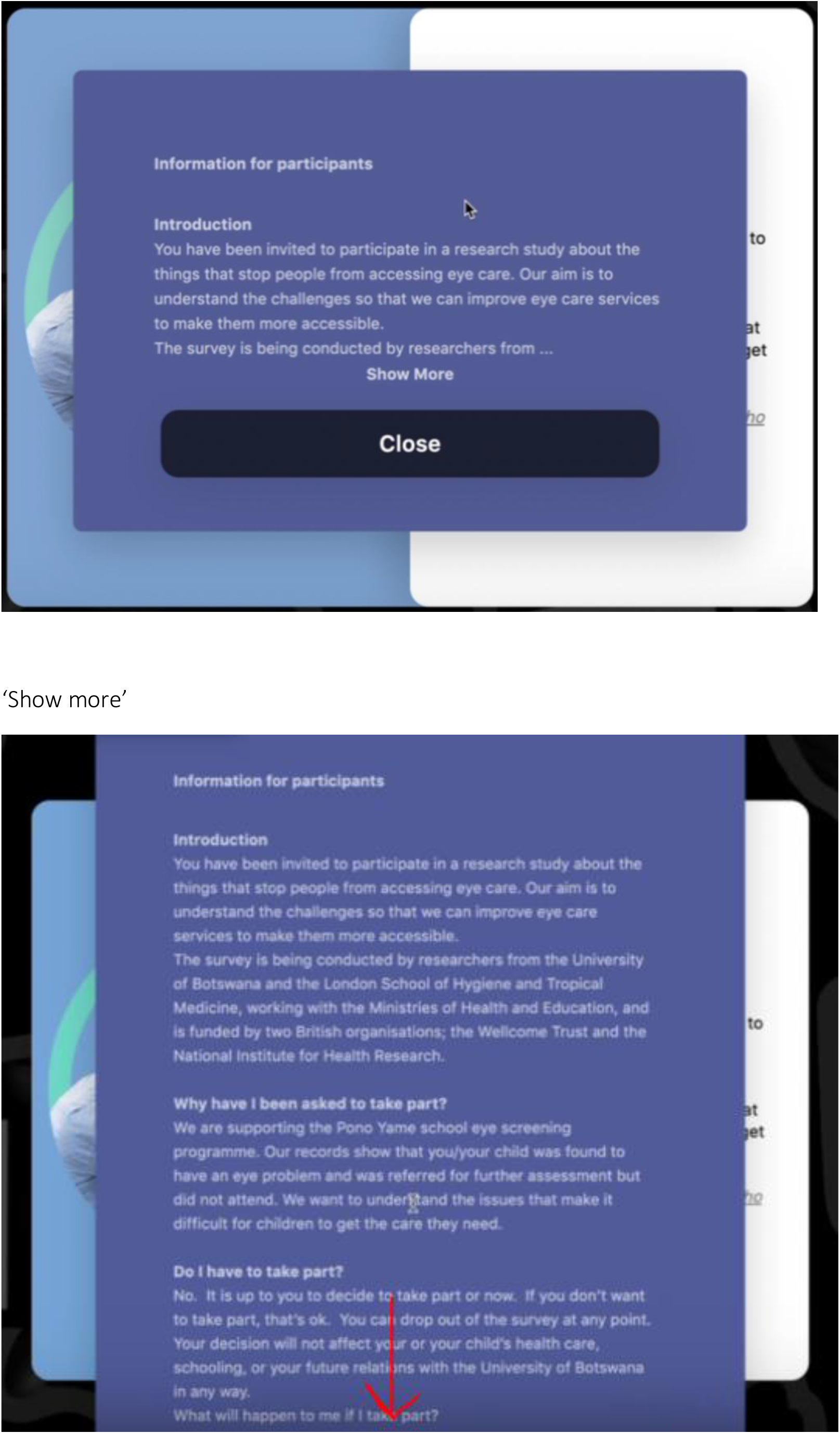

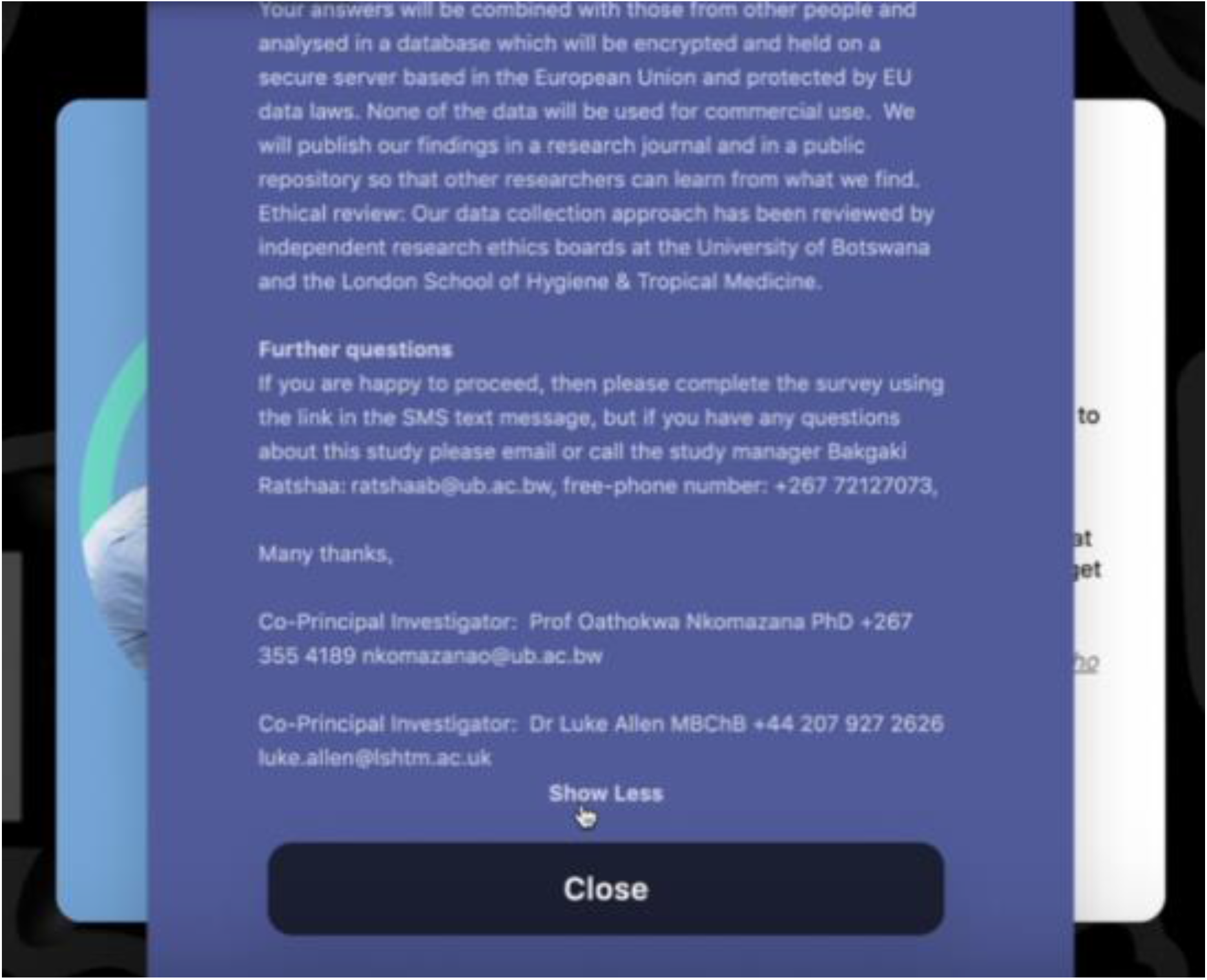

The full text is shown below:

### Participant information text for Nepal/Botswana

#### Introduction

You have been invited to participate in a research study about the things that stop people from accessing eye care. Our aim is to understand the challenges so that we can improve eye care services to make them more accessible.

The survey is being conducted by researchers from NNJS/the University of Botswana and the London School of Hygiene and Tropical Medicine, working with the Ministries of Health and Education, and is funded by two British organisations; the Wellcome Trust and the National Institute for Health Research.

#### Why have I been asked to take part?

We are supporting the Rajbiraj/Pono Yame school eye screening programme. Our records show that you/your child was found to have an eye problem and was referred for further assessment but did not attend. We want to understand the issues that make it difficult for children to get the care they need.

#### Do I have to take part?

No. It is up to you to decide to take part or now. If you don’t want to take part, that’s ok. You can drop out of the survey at any point. Your decision will not affect your or your child’s health care, schooling, or your future relations with NNJS/ the University of Botswana in any way.

#### What will happen to me if I take part?

You will complete the short online survey. It should take approximately 3 minutes.

#### Risks and benefits

Thinking about the issues that prevented you or your child from getting care may be distressing for you. There are no other risks, nor are there direct benefits to you or your child.

#### Data and confidentiality

Your /your child’s personal information will not be included and there is no way that they can be identified from any of the reports that we will produce. As the survey is anonymous we will have no way of knowing if you complete the form or not.

Your answers will be combined with those from other people and analysed in a database which will be encrypted and held on a secure server based in the European Union and protected by EU data laws. None of the data will be used for commercial use. We will publish our findings in a research journal and in a public repository so that other researchers can learn from what we find.

#### Ethical review

Our data collection approach has been reviewed by independent research ethics boards at the University of Botswana/Nepalese Ministry of Health and the London School of Hygiene & Tropical Medicine.

#### Further questions

If you are happy to proceed, then please complete the survey using the link in the SMS text message, but if you have any questions about this study please email or call the study manager Bakgaki Ratshaa, free-phone number: +267 72127073,

Many thanks,

Co-Principal Investigator: Prof Oathokwa Nkomazana PhD +267 355 4189

/Co-Principal Investigator: Dr Sailesh Mishra PhD, +977-1-4261066

Co-Principal Investigator: Dr Luke Allen MBChB +44 207 927 2626

## Appendix 2: Pilot survey questions for Botswana

| Prompt | Response type | Response |
| --- | --- | --- |
| Hi, we noticed that your child did not make it to the clinic last week. We want to understand if there is anything we could do differently to help | N/A | N/A |
| The reason my child did not attend the clinic last week is that... | Select many | 1) It's not that important to me<br>2) The clinic is hard to travel to<br>3) I can't afford to go<br>4) I can't afford the treatment<br>5) I'm worried about the treatment<br>6) My child's gender makes things difficult<br>7) The staff are rude or poorly trained<br>8) The clinic does not have enough supplies<br>9) The clinic is not open at the time I want to go<br>10) My child or I need an escort/chaperone to attend |
| Are there any other things about the eye care services or the clinic that prevented you from attending? | Freeform text | N/A |
| Would any of these changes have led to you attending? | Tap icon<br>Yes/no | 1) Help with travel<br>2) Cheaper treatment<br>3) Better explanation of what happens<br>4) Higher quality care<br>5) Different opening times<br>6) Different location<br>7) An escort or chaperone.<br>8) None of these things are enough by themselves |
| Is there anything else that we could change that would make you attend? | Select One | Yes<br>No |
| Thank you so much providing us with this feedback | Static page | N/A |

## Appendix 3: Pilot survey questions for Nepal

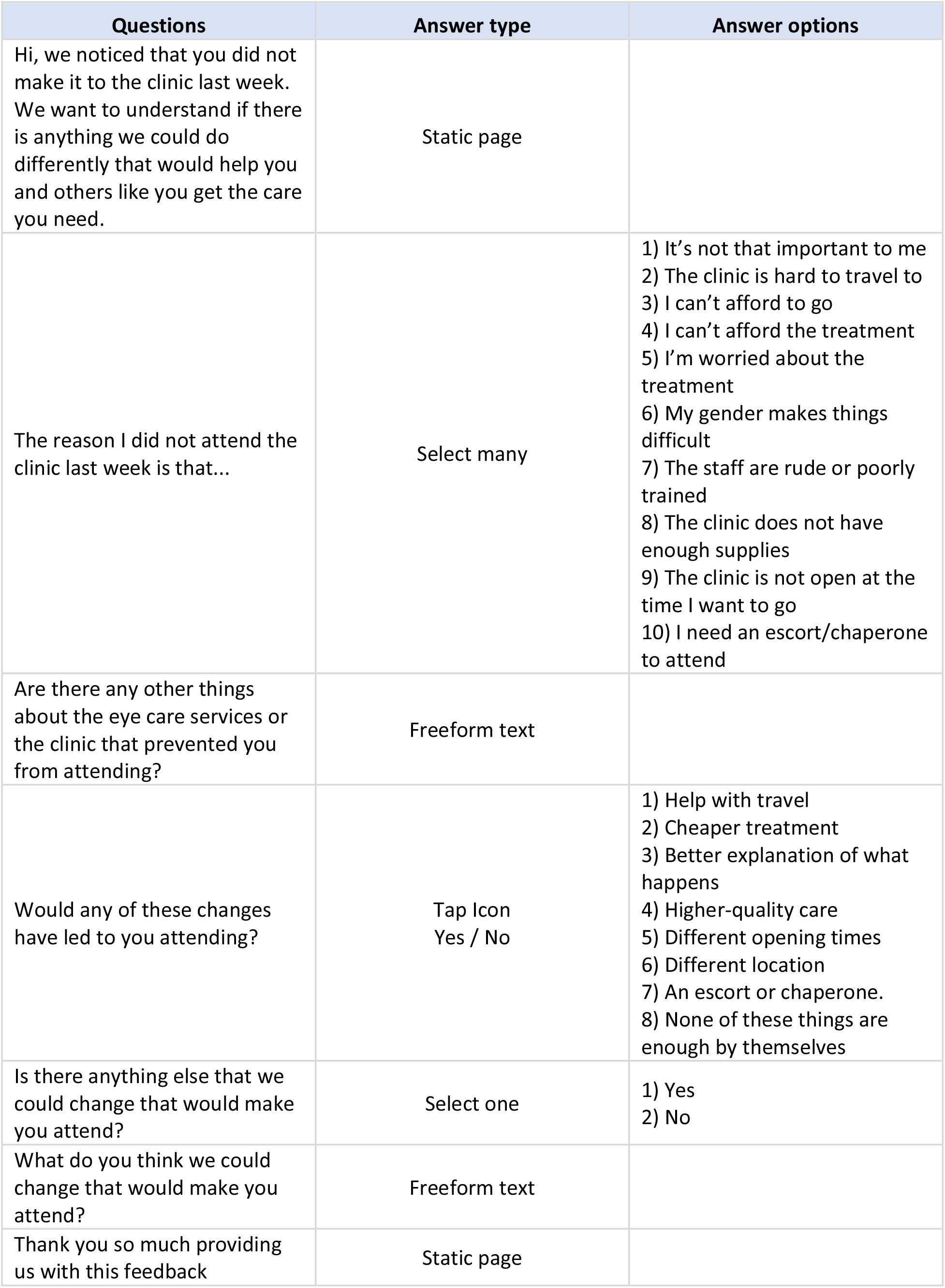

| Questions | Answer type | Answer options |
| --- | --- | --- |
| Hi, we noticed that you did not make it to the clinic last week. We want to understand if there is anything we could do differently that would help you and others like you get the care you need. | Static page |  |
| The reason I did not attend the clinic last week is that... | Select many | 1) It's not that important to me<br>2) The clinic is hard to travel to<br>3) I can't afford to go<br>4) I can't afford the treatment<br>5) I'm worried about the treatment<br>6) My gender makes things difficult<br>7) The staff are rude or poorly trained<br>8) The clinic does not have enough supplies<br>9) The clinic is not open at the time I want to go<br>10) I need an escort/chaperone to attend |
| Are there any other things about the eye care services or the clinic that prevented you from attending? | Freeform text |  |
| Would any of these changes have led to you attending? | Tap Icon<br>Yes / No | 1) Help with travel<br>2) Cheaper treatment<br>3) Better explanation of what happens<br>4) Higher-quality care<br>5) Different opening times<br>6) Different location<br>7) An escort or chaperone.<br>8) None of these things are enough by themselves |
| Is there anything else that we could change that would make you attend? | Select one | 1) Yes<br>2) No |
| What do you think we could change that would make you attend? | Freeform text |  |
| Thank you so much providing us with this feedback | Static page |  |

## Footnotes

1 University of Oxford Central University Research Ethics Committee (CUREC). Internet-mediated research. Best Practice Guidance 06_Version 7.3. Available at: https://researchsupport.web.ox.ac.uk/files/bpg-06-internet-based-research-ibr [Accessed 18/01/2023]

## Notes

FUNDERS: This work was supported by the Wellcome Trust and NIHR grant number 215633/Z/19/Z.

### Competing Interest Statement

AB is CEO of The Peek Vision Foundation and Peek Vision Ltd and receives salary support from Peek Vision. All other authors declare no competing interests.

### Funding Statement

This work was supported by the Wellcome Trust and NIHR grant number 215633/Z/19/Z.

